# Validating the Breathing Vigilance Questionnaire for use in Dysfunctional Breathing

**DOI:** 10.1101/2022.07.11.22277501

**Authors:** Jennifer Steinmann, Adam Lewis, Toby Ellmers, Mandy Jones, Vicky MacBean, Elmar Kal

## Abstract

Dysfunctional breathing (DB) is common among people with and without primary respiratory pathology. While anxiety contributes to DB, the underpinning mechanism is unclear. One explanation is that anxiety induces excessive conscious monitoring of breathing, disrupting ‘automatic’ breathing mechanics. We validated a new tool that quantifies such breathing-related ‘hypervigilance’: the Breathing Vigilance Questionnaire (Breathe-VQ).

Three-hundred-and-forty healthy adults (M_age_=27.3 years, range: 18-71; 161 men) were recruited online. We developed an initial Breathe-VQ (11 items, 1-5 Likert scale) based on the Pain Vigilance and Awareness Scale, using feedback from the target population and clinicians. At baseline, participants completed the Breathe-VQ, Nijmegen Questionnaire (NQ), State-Trait Anxiety Inventory (form 2), and Movement-Specific Reinvestment Scale (assessing general conscious processing). Eighty-three people repeated the Breathe-VQ two weeks later.

We removed five items based on item-level analysis. The resulting six-item Breathe-VQ questionnaire (score range: 6-30) has excellent internal (alpha=.892) and test-retest reliability (ICC=.810), a minimal detectable change of 6.5, and no floor/ceiling effects. Concurrent validity was evidenced by significant positive correlations with trait anxiety and conscious processing scores (*r*’s=.35-.46). Participants at high-risk of having DB (NQ>23; N=76) had significantly higher Breathe-VQ score (M=19.1±5.0) than low-risk peers (N=225; M=13.8±5.4; *p*<.001). In this ‘high-risk’ group, Breathe-VQ and NQ-scores were significantly associated (*p*=.005), even when controlling for risk factors (e.g., trait anxiety).

The Breathe-VQ is a valid and reliable tool to measure breathing vigilance. Breathing vigilance may contribute to DB, and could represent a therapeutic target. Further research is warranted to further test the Breathe-VQ’s prognostic value, and assess intervention effects.

**Key Findings:**

– Dysfunctional breathing (DB) is highly prevalent in the general population as well as in people with respiratory conditions.
– Anxiety is identified as a key factor contributing to DB, potentially because it induces conscious, anxious monitoring of breathing.
– We developed a short self-reported outcome measure of such breathing-specific vigilance, the Breathe-VQ.
– The Breathe-VQ was found to be a valid and reliable tool for use in the general population.
– Breathe-VQ scores were positively associated with self-reported breathing problems, after correcting for known risk factors such as trait-anxiety.

## 1. Introduction

Dysfunctional breathing (DB) is a breathing disorder where people demonstrate maladaptive breathing pattern changes, such as hyperventilation [1,2], erratic breathing [2,3], reduced breath holding ability [4], and frequent sighing [5]. People with dysfunctional breathing (PWDB) frequently experience air hunger, in addition to non-breathing related symptoms (e.g., pain, dizziness; [6]), and report reduced quality of life [3,7]. DB frequently occurs *secondary* to specific respiratory conditions, such as asthma and Chronic Obstructive Pulmonary Disease (COPD; [8]), and affects many people with ‘long COVID’ [9]. However, for around 10-20% of the general population, DB is *primary* [10,11], and cannot be linked to clear pathophysiological changes [2].

Breathing exercises are a primary component of treatment of DB [1,12]. Such exercises are intended to ‘retrain’ breathing control, enabling individuals to shift toward diaphragmatic breathing, lower respiratory rate, and reduce upper-chest excursions while breathing [1,12]. Usually these breathing exercises are accompanied by education on DB and relaxation techniques [13], as DB seems to be linked to anxiety and associated changes in attention [14,15]. However, whilst some studies show promising results [13,15], there is currently no conclusive evidence for any specific treatment of DB [12].

One factor that complicates the treatment of DB is that its aetiology is often unclear. Psychological factors, especially anxiety, may directly alter breathing control [16], and play a key role in the onset and maintenance of DB symptoms [15,17,18]. In line with Vidotto et al. [14], we argue that anxiety-related disruptions of interoceptive awareness may contribute to DB. Interoception is “…the ability to identify, access, understand, and respond appropriately to the patterns of internal signals” (p3 [19], [20]). Several studies show disturbed interoceptive processing of breathing in high-anxious individuals [22]. Specifically, in PWDB there seems to be anxiety-induced excessive monitoring of breathing [14], making them more likely to notice breathing changes and interpret these as threatening (even if innocuous). This may result in a vicious cycle, whereby attempts at consciously controlling breathing lead to maladaptive respiratory alterations, which in turn reinforce anxiety and vigilance [21]. Such anxiety related ‘hypervigilance’ towards interoceptive bodily signals has also been implicated in a variety of other disorders that lack a clear neuro-biological basis [23-26].

To investigate the role of breathing-related hypervigilance in DB, we need a validated outcome measure that can reliably assess this. Several measurement instruments exist that investigate related constructs, such as the Breathlessness Beliefs Questionnaire (BBQ; [27]), the Multidimensional Dyspnoea Profile (MDP; [28]), and the Dyspnoea-12 [29]. However, none measure *hypervigilance* directly, but rather its (indirect) influence on e.g. beliefs about breathing symptoms. The Multidimensional Assessment of Interoceptive Awareness (MAIA) questionnaire [30] and Body Vigilance Scale [21] both combine concepts of awareness of bodily sensations and different factors relating to attention, but neither were developed specifically for breathing – which limits their utility for use in DB, as hypervigilance is likely domain-specific [31].

Therefore, the current study aimed to develop and validate a self-reported breathing-specific vigilance questionnaire (Breathe-VQ) that directly measures (hyper)vigilance of breathing, and captures the potential interplay between conscious monitoring/control of breathing and anxiety. For this purpose, we adapted a pain-specific measure (the Pain Vigilance and Awareness Questionnaire; [23]) and validated the resulting Breathe-VQ in a large sample recruited from the general population, in which primary DB is known to be prevalent [10,11].

## 2. Methods

### 2.1. Participants

#### 2.1.1. Recruitment

Three-hundred-and-forty healthy adults were recruited for this study. Regarding sample size, key analyses in this study were the factor analysis and retest reliability analysis (see section 2.4). For the former, a subject-to-variable ratio of at least 10:1 has been recommended, but we erred on side of caution and aimed for two samples of 150-200 participants for each analysis [32]. For test-retest reliability, we aimed to have a minimal number of 60 individuals with complete data for the Breathe-VQ at both T1 and T2, as this would ensure 80% power to detect an intraclass correlation coefficient of .80 (95%CI: .70-.90). Anticipating drop-out, we invited the first 130 participants for T2, but stopped once 90 participants had completed the questionnaire at T2.

Recruitment took place online, using two complementary modes of recruitment: (i) Recruitment through Brunel University London’s Division of Psychology Research Participant Sign-up System (SONA); (ii) Recruitment through ‘Testable Minds’ (https://www.testable.org/), a GDPR-compliant, well-established global online platform for participant recruitment. Participants recruited through SONA were given study credits in exchange for participation, while participants recruited through Testable Minds were given monetary compensation ($3).

Participants were eligible for inclusion if they (i) were ≥18 years of age, (ii) had no self-reported diagnosis of respiratory and/or cardiac conditions, (iii) had no diagnosis of COVID-19 within the preceding three months and/or chronic COVID syndrome (“long-COVID”).^1^

Institutional ethical approval was obtained from the College of Health, Medicine and Life Sciences Research Ethics Committee of Brunel University London. All participants provided online written informed consent prior to participation.

### 2.2. Measurement instruments

#### 2.2.1. Breathe-VQ – Initial development

The Breathe-VQ was created by adapting the adult and children versions of the Pain Vigilance and Awareness Scale [23,33]. Researchers with expertise in respiratory research and/or psychological theory (JS, EK, TE, VM) created an initial Breathe-VQ version. Other members of the research team (MJ, AL; respiratory physiotherapists) gave feedback on this version, and their relevance to (dysfunctional) breathing, after which the Breathe-VQ was adapted accordingly. An Open Science Framework page (**https://osf.io/shqtf/**) details the (justification for) different iterations and changes made. The final agreed-upon Breathe-VQ that was completed by participants for further validation is presented in Table 1.

**Table 1.** Initial 11-item version of the Breathe-VQ.

|  | Never |  | Sometimes |  | Always |
| --- | --- | --- | --- | --- | --- |
| 1. I closely monitor how difficult my breathing feels | 1 | 2 | 3 | 4 | 5 |
| 2. I become alarmed when I experience breathlessness or tightness in my chest | 1 | 2 | 3 | 4 | 5 |
| 3. I am highly aware of small changes in how my breathing feels | 1 | 2 | 3 | 4 | 5 |
| 4. I feel as if I am more aware of my breathing than other people | 1 | 2 | 3 | 4 | 5 |
| 5. When something happens that affects my breathing, I am anxious to work out how breathless I am | 1 | 2 | 3 | 4 | 5 |
| 6. I worry about fluctuations in my breathing | 1 | 2 | 3 | 4 | 5 |
| 7. I avoid situations that I fear will increase feelings of breathlessness | 1 | 2 | 3 | 4 | 5 |
| 8. I become preoccupied with monitoring my breathing | 1 | 2 | 3 | 4 | 5 |
| 9. I remain calm in situations that affect my breathing | 1 | 2 | 3 | 4 | 5 |
| 10. I worry that physical activity will increase my sensation of breathlessness | 1 | 2 | 3 | 4 | 5 |
| 11. I dwell on my breathing | 1 | 2 | 3 | 4 | 5 |

**NB:** Instructions were as follows: “Please read the sentences below and choose a number between 1 (never) and 5 (always) that best describes how you typically feel in relation to your breathing.”

#### 2.2.2. Nijmegen Questionnaire

We used the Nijmegen Questionnaire (NQ; [34]) to screen symptoms indicative of dysfunctional breathing. This measure comprises 16 items (scores 0-4; total score: 0-64). Scores >23 suggest hyperventilation syndrome, a type of dysfunctional breathing [34].

#### 2.2.3. Trait anxiety and movement-specific reinvestment

For the concurrent validity analysis, we assessed both trait-anxiety and trait-propensity to focus on movement.

We assessed trait-anxiety using the State-Trait Anxiety Inventory (STAI-2; [35]). The Trait form contains 20 items (scored 1-4), and total scores range between 0-80. Higher scores indicate greater trait anxiety.

The Movement-Specific Reinvestment Scale (MSRS; [36]) measured how much people consciously attend to their movements. This questionnaire contains 10 items, scored from one (“strongly disagree”) to six (“strongly agree”). Five items form the subscale “Conscious Motor Processing” (probing *control* of movement), while the other 5 items form the “Movement Self-Consciousness” subscale (probing movement self-awareness). Subscale scores range from 5-30, higher scores reflecting greater conscious movement processing.

### 2.3. Procedures

#### 2.3.1. Timepoint 1 (T1)

Participants completed the study online. After providing informed consent, participants answered screening questions, to determine eligibility. They would then complete additional questions on age, sex, general health, (earlier) diagnosis of anxiety and/or depression, followed by the Breathe-VQ, NQ, MSRS, and STAI-2 (in this order).

#### 2.3.2. Timepoint 2 (T2)

To assess test-retest reliability, participants received an email invitation to complete the Breathe-VQ a second time, two weeks after T1 (M: 14.7±2.7, range: 13-26). If necessary, a one-off reminder email was sent one week later. This time period was considered sufficient to minimise recall bias.

### 2.4. Data analysis and statistics

All data were analysed with SPSS and AMOS (version 26; IBM, Chicago, IL). Alpha was set at *p*<.05. Figure 1 summarises the flow of the study and analyses. Analyses involved four different steps:

**Figure 1.**
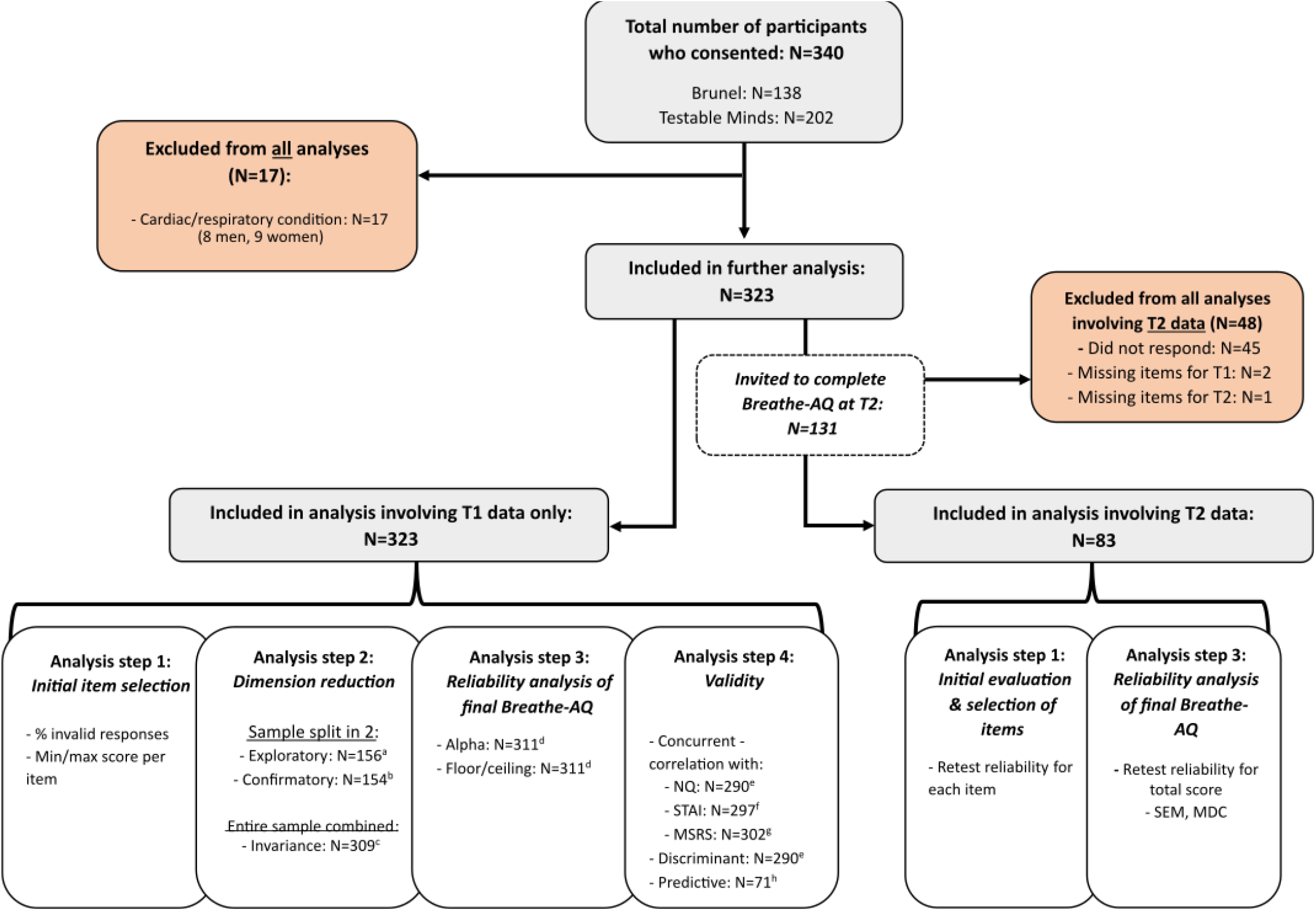
Study flow. Participants were recruited (online) through Brunel and Testable Minds. The figure shows who were in- and excluded for which analysis, and why. ^a^ 8 participants excluded (missing value(s)); ^b^ 5 participants excluded (missing value(s)); ^c^ 14 participants excluded (N=2: stated they did not identify as female/male; N=12: missing values); ^d^ 12 participants excluded (missing values); ^e^ 33 participants excluded (N=21: missing value for NQ; N=1: missing value for NQ & Breathe-VQ; N=11: missing value for Breathe-VQ); ^f^ 26 participants excluded (N=14: missing value for STAI; N=4 missing value for both STAI & Breathe-VQ; N=8: missing value for Breathe-VQ); ^g^ 21 participants excluded (N=9: missing value for MSRS; N=2 missing value for both MSRS & Breathe-VQ; N=10: missing value for Breathe-VQ); ^h^ 76 participants initially included, as their NQ scores >23. 5 of these excluded due to missing STAI or Breathe-VQ scores;

#### 2.4.1. Step 1 – Initial screening of items

In step 1, we screened individual items’ behaviour. We flagged items for which:

- there were a large number of missing (or multiple) responses (>5%)
- >50% of responses were the minimum or maximum score
- for which test-retest reliability was low (2-way, random effect, consistency single measures ICC<.5; [37]).

The research team discussed flagged items, and reached agreement on whether these should be excluded from the subsequent analysis steps.

#### 2.4.2. Step 2 - Dimension reduction and validation

Step 2 concerned exploratory principal component analysis and subsequent confirmatory factor analysis. Participants were first randomly allocated (using random.org, 50:50 ratio) to either an ‘exploratory’ or ‘confirmatory’ subsample (see Figure 1). Exploratory analysis (varimax rotation) was done using the T1 Breathe-VQ data (on items retained after step 1). The inflection point in the scree plot was used to identify the number of components of the scale. We considered removal of items that loaded insufficiently (<0.4; [38]) on a component, loaded on more than one component, and/or showed low item-rest correlations (r<0.3).

Next, confirmatory factor analysis was performed to assess if the data fitted the component-structure as determined with the preceding exploratory analysis. We used the T1 data of the ‘confirmatory’ subgroup. The procedure entailed analysis of the variance-covariance matrix with maximum likelihood estimation [39]. Items were constrained to load on the component(s) they should load on based on the exploratory principal component analysis. Pairs of error terms within each factor were allowed to co-vary if this improved model fit. Model fit was evaluated using standard criteria (see Supplementary material 3 for details [40-42]).

Subsequently, “measurement invariance” was determined, to assess whether the scale structure was similar for men and women – this because women are more likely to experience DB [10], which may affect their interpretation of the questionnaire. See Supplementary material 3 for details [43].

#### 2.4.3. Step 3 - Reliability and measurement error

We assessed internal consistency (Cronbach’s alpha) and test-retest reliability (2-way, random effect, consistency, single measures ICC) of the finalised Breathe-VQ. Alpha and ICC >.70 indicate sufficient reliability. We further determined measurement error (SEM = SD + 2*√(1-ICC); [44]), and minimal detectable change on group and individual level (MDC_group_= SEM × 1.96 × √2/√n; MDC_individual_ = SEM × 1.96 × √2; [45]). Finally, we screened for floor and ceiling effects for the total Breathe-VQ score (i.e., >15% of participants scoring lowest/highest possible scores [46,47]).

#### 2.4.4. Step 4 - Concurrent and discriminant validity

Concurrent validity was assessed by correlating (Pearson’s *r*) Breathe-VQ total scores with (i) STAI, and (ii) MSRS subscale scores.

To assess discriminant validity, we used independent samples t-test to assess whether people at risk of having DB (NQ>23) have higher total Breathe-VQ scores compared to low-risk peers (NQ≤23).

Finally, linear regression analysis investigated whether total Breathe-VQ scores would be significantly associated with severity of DB-related symptoms (NQ) *within the group of people at risk of DB* (see above), when controlling for confounding variables (age, gender, trait-anxiety score, and depression diagnosis; [10,14,15,17]).

## 3. Results

### 3.1. Participant characteristics

Figure 1 summarises the flow of the study. In total, 340 participants completed the study at T1, of which 17 were excluded due to self-reported respiratory and/or cardiovascular diagnosis. Table 2 lists the characteristics of the remaining 323 participants. Participants were relatively young and scored relatively high on the Nijmegen Questionnaire and STAI-2. Supplementary material 1 summarises the characteristics of the test-retest sample (i.e., those individuals who also completed the questionnaire at T2). This subsample was found to be somewhat younger, to include more women, and to have a higher score on the NQ compared to the overall sample.

**Table 2.** Characteristics of total sample (N=323).

|  |  |  |
| --- | --- | --- |
| **General** | Male / Female / Non-binary (N) | 161 / 160 / 2 |
| | Age (years; M ±SD (range)) | 27.3 ± 9.8 (18–71)<sup>a</sup> |
| **Nijmegen Questionnaire** | Total score (M ±SD (range)) | 17.8 ± 10.0 (0–49)<sup>b</sup> |
|  | Score>23 (n, %) | 76 (24%) |
| **Self-reported General Health** | Excellent (n (%)) | 74 (22.9%) |
|  | Very Good (n (%)) | 142 (44.0%) |
|  | Good (n (%)) | 85 (26.3%) |
|  | Fair (n (%)) | 16 (5.0%) |
|  | Poor (n (%)) | 3 (0.9%) |
|  | Missing (n (%)) | 3 (0.9%) |
| **Psychological Characteristics / Traits** | Diagnosis of Depression (n (%)) | 51 (16%) |
|  | Diagnosis of Anxiety (n (%)) | 68 (21%) |
| | Trait Anxiety (STAI-2; M ±SD (range)) | 46.6 ± 12.4 (21–80)<sup>c</sup> |
| | MSRS – CMP (M ±SD (range)) | 15.9 ± 5.7 (5–30)<sup>d</sup> |
| | MSRS – MS-C (M ±SD (range)) | 16.2 ± 6.7 (5–30)<sup>d</sup> |

<sup>a</sup>22 missing values; <sup>b</sup>1 missing value; <sup>c</sup>18 missing values; <sup>d</sup>11 missing values;
**Abbreviations:** M = mean; MSRS – CMP = Movement-Specific Reinvestment Scale, Conscious Movement Processing subscale; MSRS – MS-C = Movement-Specific Reinvestment Scale, Movement Self-Consciousness subscale; n = number; SD = standard deviation; STAI-2 = State-Trait Anxiety form 2 (trait assessment);

### 3.2 Step 1 – Initial screening of items

For the initial 11-item Breathe-VQ, no clear issues were noted regarding missing values (N=26 in total, N≤6 (1.9%) for separate items). Reliability was acceptable to good for items 1-6 and 10-11 (ICC≥.581, range: .581-.704). Items 7 (ICC=.466) and 9 (ICC=.329) had low test-retest reliability (ICC<.500). Item 8 showed a potential floor effect (minimum value >50% of responses). Therefore, items 7-9 were removed from the questionnaire prior to further analyses. Supplementary material 2 summarises item-level characteristics.

### 3.3. Step 2 - Dimension reduction and validation

#### 3.3.1. Principal component analysis

Principal component analysis on the 8 selected items (items 1-6, and items 10-11) revealed a two-component solution (Table 3). Only items 10 and 11 were linked to component 2. Upon reflection, we deemed item 10 to not fully capture breathing vigilance, but rather behavioural consequences. Item 11’s substantial loading on both components (Table 3) suggests issues with this item’s interpretation. Coupled to their borderline floor effect (42% and 46%, see Supplementary material 3) we thus decided to remove these items, and run the analysis a second time. As shown in Table 3, now all six items loaded highly on one component only. Items 1-6 were therefore selected for the subsequent confirmatory factor analysis.

**Table 3.** Component loadings for each item, presented separately for each of the two runs of the principal component analysis.

| Item | RUN 1 <sup>a</sup> |  | RUN 2 <sup>b</sup><br>(after excluding items 10, 11) |
| --- | --- | --- | --- |
|  | Component Loading<br>(explained variance 72.0%) |  | Component Loading<br>(explained variance 68.8%) |
|  | *Component 1* | *Component 2* | *Component 1* |
| 1. I closely monitor how difficult my breathing feels | **.784** | .288 | **.833** |
| 2. I become alarmed when I experience breathlessness or tightness in my chest | **.776** | .267 | **.832** |
| 3. I am highly aware of small changes in how my breathing feels | **.860** | -.030 | **.813** |
| 4. I feel as if I am more aware of my breathing than other people | **.802** | .174 | **.813** |
| 5. When something happens that affects my breathing, I am anxious to work out how breathless I am | **.810** | .235 | **.849** |
| 6. I worry about fluctuations in my breathing | **.783** | .297 | **.837** |
| 10. I worry that physical activity will increase my sensation of breathlessness | .071 | **.941** | *n/a* |
| 11. I dwell on my breathing | .529 | **.628** | *n/a* |

<sup>a</sup> Kaiser-Meyer-Olkin assessment (KMO)=.899; all individual KMOs≥.748 (>0.5 threshold [32]).
<sup>b</sup> KMO=.900; individual KMOs≥.890;

#### 3.3.2. Confirmatory factor analysis

Item-factor loadings were positive and high (.64-.81), and model fit indices were good (*χ*^2^(8)=10.046, *p*=.262; *χ*^2^/df=1.256; CFI=.995; GFI=.978; RMSEA=.041 [.000, .108]; SRMR=0.030). Further tests supported measurement invariance, which indicates that the scale structure is similar across men and women. See Supplemental material 3 for further details.

### 3.4. Step 3 - Reliability and measurement error

The final Breathe-VQ is presented in Figure 2.

**Figure 2.**
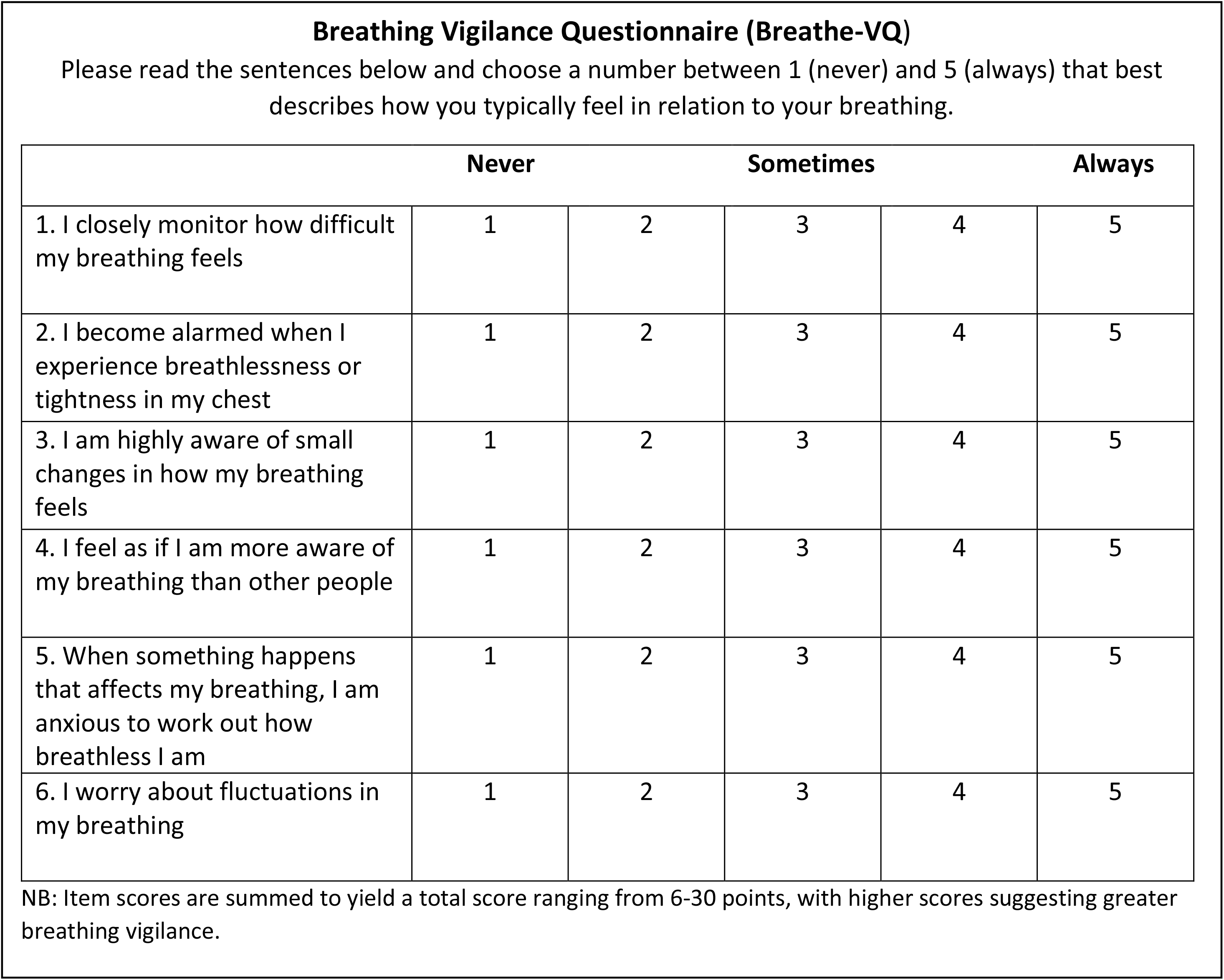
Final Breathing Vigilance Questionnaire (Breathe-VQ).

The test-retest sample’s (N=83; Figure 1) Breathe-VQ data showed excellent retest-reliability (ICC=.810, 95%CI[.721, .873]). Standard error of measurement was 2.33 points. As such, the minimal detectable change was estimated at 0.7 on group level, and 6.5 on individual level.

We found excellent internal consistency (alpha = .892). No indications of floor or ceiling effects were evident, as only 5.0% (N=16) of individuals scored the minimal possible score (6 points), and 1.2% (N=4) scored the maximal possible score (30 points).

### 3.5. Step 4 - Validity

Regarding concurrent validity, Breathe-VQ sum scores significantly correlated to scores on the STAI (*r*=.351, *p*<.001, N=297), and participants’ Conscious Motor Processing (*r*=.459, *p*<.001, N=302) and Movement Self-Consciousness (*r*=.385, *p*<.001, N=302) scores.

Regarding discriminant validity, the ‘low risk of DB’ group (NQ<24; N=216) had significantly lower scores (M=13.8, SD=5.4) on the Breathe-VQ compared to the 74 people in the ‘high risk of DB’ group (M=19.1, SD=5.0; *t*(288)=7.760, *p*<.001).

Finally, linear regression analysis showed that, within the ‘high risk of DB’ group, Breathe-VQ scores were significantly associated with the scores on the NQ – even when controlling for confounding variables (trait anxiety, age, sex, depression diagnosis). That is, explained variance significantly increased when Breathe-VQ scores were added in a second analysis step (ΔR^2^=.100, *p*=.005; see supplementary material 4).

## 4. Discussion

This study describes the development of the novel, simple-to-use Breathe-VQ. This is a self-reported outcome measure of an individual’s conscious monitoring of their breathing state. We show the questionnaire to be valid and reliable, and provide minimal important differences at group and individual level. The Breathe-VQ is a very simple six-question patient-reported questionnaire which would be quick to administer in clinical practice. It performs well without floor and ceiling effects and now has established minimal differences. Further, Breathe-VQ scores are positively associated with NQ scores in a sample of participants at risk of having DB. This suggests that the questionnaire could have clinical utility for predicting DB (severity) in the general population.

Breathing is typically a mostly automated physiological function that requires little conscious monitoring or control. However, in our sample, many participants at risk of DB display vigilant monitoring of their breathing. While we cannot draw causal inferences based on our cross-sectional data, there is a real likelihood that this vigilance may in fact be excessive (i.e., they may be “hypervigilant” toward breathing), and contributes to and/or helps maintain breathing related complaints. Studies on balance control, which like breathing is traditionally viewed as an ‘automatic’ physiological function, show that people will become consciously focused on their balance during situations that threaten their stability (e.g., walking across uneven ground or standing at height). This, in turn, has been shown to induce distorted perceptions of instability – whereby people perceive themselves to be more imbalanced than they actually are [26]. It seems plausible that the same mechanisms may be at play in people with DB. Note though, that in the current study, the greater breathing vigilance reported by people at risk of DB may also be *the result of* having experienced maladaptive breathing. Likely, a reciprocal relationship exists, where hypervigilance may both be triggered by, and a trigger of, disrupted breathing mechanics. Future studies may further look into this.

The Breathe-VQ provides a means to screen for breathing-specific vigilance, and could have clear clinical use. For people with excessive breathing-related vigilance, it may be useful to adopt treatment methods that aim to help ‘recalibrate’ perceptions and appraisal of breathing ([49]). Mindfulness based approaches may help in this regard [49], especially in combination with exercises aimed at re-educating interpretation of breathing related bodily signals, and anxiety-alleviating interventions. Some arts-in-health practices such as Singing for Lung Health [50] may be useful in this regard, as well as more generally used mind-body movement therapies such as yoga, or tai-chi [49].

### Limitations

Data were collected during a period in which there were very strict COVID-19 restrictions. As such, participants may have been more relatively more aware of their breathing in general. Indeed, this may explain the relatively high proportion of people with elevated trait anxiety and NQ scores in our sample. Second, we used a threshold of greater than 23 on the NQ and, while this may indicate a greater risk of having DB, it is not by itself sufficient to diagnose DB. Third, there were differences in age and gender between the overall sample and the subsample who repeated the questionnaire completion for test-retest reliability purposes. Yet, as the confirmatory factor analysis revealed measurement invariance for gender, we are confident this did not substantially influence our results.

### Further research

Further work is now needed to investigate if the questionnaire scores can be used to predict future development of DB, and/or changes in DB severity over time. This would require studies in which the questionnaire is tested in a sample who have confirmed DB (diagnosed by a trained clinician, using appropriate multidimensional assessment methods (51)). The questionnaire should also be tested in people who have chronic respiratory diseases such as Asthma, COPD, Interstitial Lung Disease and Bronchiectasis, and determine its responsiveness to change following pulmonary rehabilitation.

## Conclusion

Breathlessness and dysfunctional breathing in the absence of clear underlying pathology is a very common health issue with incompletely understood underpinning mechanisms. We adapted a pain vigilance questionnaire to develop a breathing specific vigilance questionnaire. This Breathe-VQ is a valid and reliable tool to measure vigilance of breathing in an otherwise healthy population consisting of individuals with and without suspected DB. Further research is now warranted exploring the Breathe-VQ in clinical populations and establishing intervention effects on vigilance of breathing.

## Rights Retention Strategy Statement

This research was supported by Brunel University London, publicly funded by Research England. A CC BY is applied to the AAM arising from this submission, in accordance with the University’s Open Access Mandate.

## Data Availability

All data produced in this study are available in the manuscript, supplementary data and via an Open Science Framework page (https://osf.io/shqtf/)

## Supplemental Material 1. Characteristics of test-retest subsample

**Table S1.**
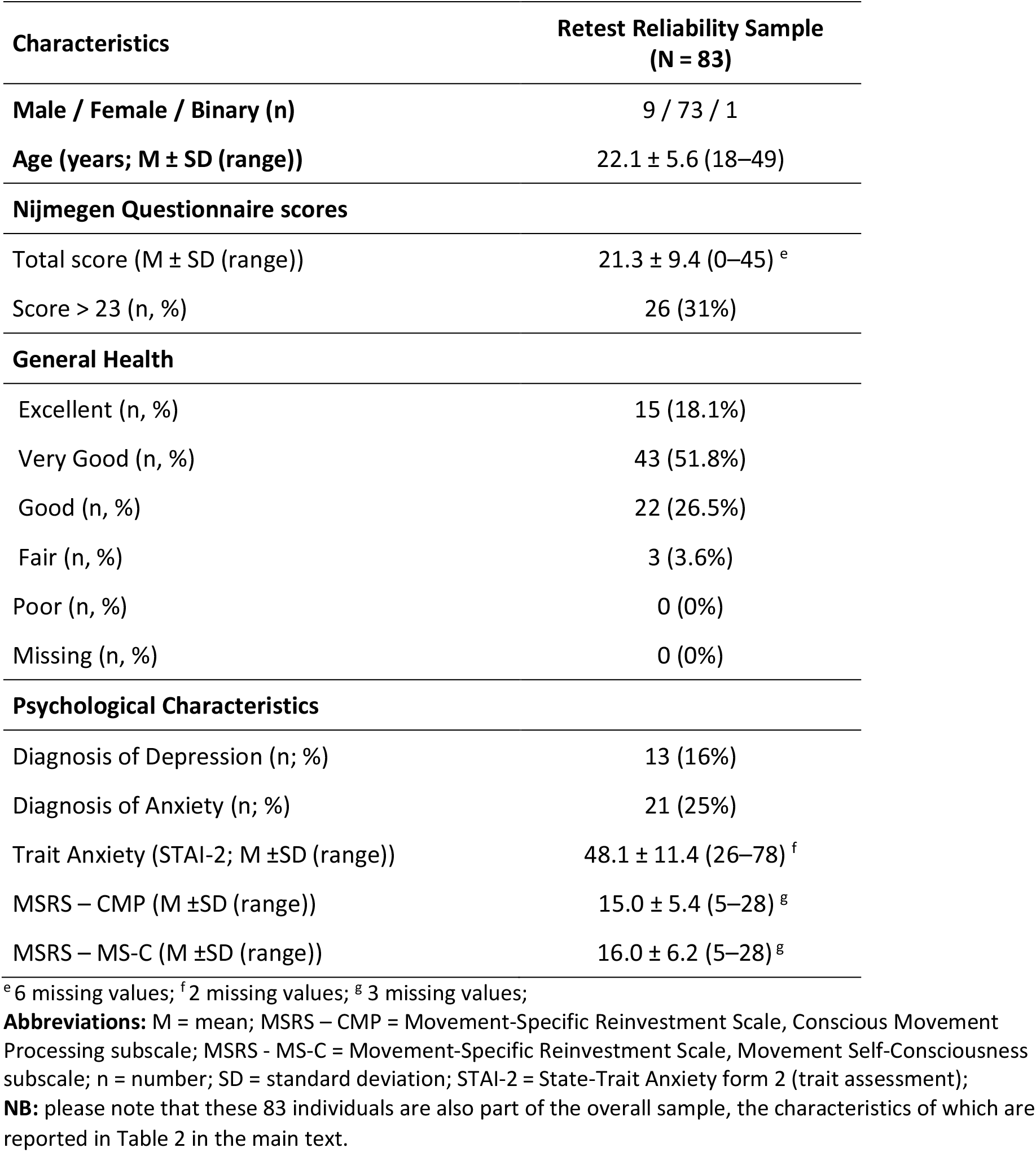

| Characteristics | Retest Reliability Sample<br>(N = 83) |
| --- | --- |
| Male / Female / Binary (n) | 9 / 73 / 1 |
| Age (years; M ± SD (range)) | 22.1 ± 5.6 (18–49) |
| **Nijmegen Questionnaire scores** |  |
| Total score (M ± SD (range)) | 21.3 ± 9.4 (0–45) <sup>e</sup> |
| Score > 23 (n, %) | 26 (31%) |
| **General Health** |  |
| Excellent (n, %) | 15 (18.1%) |
| Very Good (n, %) | 43 (51.8%) |
| Good (n, %) | 22 (26.5%) |
| Fair (n, %) | 3 (3.6%) |
| Poor (n, %) | 0 (0%) |
| Missing (n, %) | 0 (0%) |
| **Psychological Characteristics** |  |
| Diagnosis of Depression (n; %) | 13 (16%) |
| Diagnosis of Anxiety (n; %) | 21 (25%) |
| Trait Anxiety (STAI-2; M ±SD (range)) | 48.1 ± 11.4 (26–78) <sup>f</sup> |
| MSRS – CMP (M ±SD (range)) | 15.0 ± 5.4 (5–28) <sup>g</sup> |
| MSRS – MS-C (M ±SD (range)) | 16.0 ± 6.2 (5–28) <sup>g</sup> |

<sup>e</sup> 6 missing values; <sup>f</sup> 2 missing values; <sup>g</sup> 3 missing values;
**Abbreviations:** M = mean; MSRS – CMP = Movement-Specific Reinvestment Scale, Conscious Movement Processing subscale; MSRS - MS-C = Movement-Specific Reinvestment Scale, Movement Self-Consciousness subscale; n = number; SD = standard deviation; STAI-2 = State-Trait Anxiety form 2 (trait assessment);

## Supplementary Material 2. Results of initial screening of items

**Table S2.**

| *Item* | *n / %*<br>*missing* | *% min/max*<br>*score* | *ICC (95% CI)* | *Included?* |
| --- | --- | --- | --- | --- |
| 1. I closely monitor how difficult my breathing feels | 2 / 0.6% | 27% / 4% | .705 (.577, .799) | Yes |
| 2. I become alarmed when I experience breathlessness or tightness in my chest | 2 / 0.6% | 16% / 16% | .573 (.409, .702) | Yes |
| 3. I am highly aware of small changes in how my breathing feels | 1 / 0.3% | 20% / 7% | .609 (.454, .728) | Yes |
| 4. I feel as if I am more aware of my breathing than other people | 2 / 0.6% | 34% / 6% | .705 (.578, .799) | Yes |
| 5. When something happens that affects my breathing, I am anxious to work out how breathless I am | 1 / 0.3% | 25% / 9% | .692 (.561, .790) | Yes |
| 6. I worry about fluctuations in my breathing | 4 / 1.2% | 35% / 4% | .646 (.500, .755) | Yes |
| 7. I avoid situations that I fear will increase feelings of breathlessness | 3 / 0.9% | 43% / 6% | .464 (.277, .617) | No |
| 8. I become preoccupied with monitoring my breathing | 2 / 0.6% | 54% / 1% | .571 (.406, .701) | No |
| 9. I remain calm in situations that affect my breathing | 6 / 1.9% | 7% / 14% | .381 (.181, .550) | No |
| 10. I worry that physical activity will increase my sensation of breathlessness | 2 / 0.6% | 42% / 4% | .712 (.588, .804) | Yes |
| 11. I dwell on my breathing | 1 / 0.3% | 46% / 1% | .675 (.538, .777) | Yes |

**NB:** Predetermined cut-off values were 5% (missing cases per item), 50% (% of maximal / minimal scores for an item), and ICC<.500. Excluded items – items 7, 8, and 9 - are highlighted in red.

## Supplementary Material 3. Factor Analyses

For the confirmatory factor analysis, we evaluated model fit of a model where items 1-6 were constrained to load on one underlying factor/construct (based on the exploratory analysis’ results). T1 data from the ‘confirmatory subsample’ were used for this purpose. We then assessed the standardised item-factor loadings, the chi-square statistic – both raw (χ^2^) and divided by its degrees of freedom (χ^2^/df; both should be close to zero for good fit), goodness-of-fit and comparative fit indices (CFI; values>.95 indicate good fit), standardized root mean squared residual (SRMR; values<.08 indicate good fit), and the root mean square error of approximation (RMSEA; values<.05 indicate good fit [40-42].

In an initial run, we found standardised item-factor loadings for items 1-6 to be positive and high (.65-.79). While model fit indices showed mixed results (*χ*^*2*^(9)=26.338, *p*=.002; *χ*^*2*^/df=2.926; CFI=.958; GFI=.941; RMSEA=.112 [.064, .163]; SRMR=0.043), inspection of modification indices revealed model fit could be improved by allowing items 5 and 6’s error terms to covary (MI=12.584). In a second analysis run, we found that item-factor loadings remained positive and high when these error terms covaried (.64-.81; Figure S3). Further, model fit indices substantially improved, and were now good overall: *χ*^2^(8)=10.046, *p*=.262; *χ*^2^/df=1.256; CFI=.995; GFI=.978; RMSEA=.041 [.000, .108]; SRMR=0.030.

Table S3 shows the results of measurement invariance testing. For this analysis, model fit was assessed when item-factor loadings were free to differ between male and female subgroups (configural invariance), when item-factor loadings were equated across groups (so-called metric invariance testing), and when both the item-factor loadings and the intercepts of the model were equated across groups (so-called scalar invariance). As model fit remained statistically similar across all these three steps – i.e., non-significant change in χ^2^, ΔCFI<0.010 ΔRMSEA<0.015, and ΔSRMR<0.030 (metric invariance) or <0.010 (scalar invariance) – the scale’s structure can be considered to be similar regardless of group status (cut-offs based on [43]).

In sum, confirmatory factor analysis supported the results obtained by the exploratory principal component analysis: We can be confident the scale taps into one underlying construct (breathing vigilance) and that this scale structure is similar for men and women (measurement invariance).

**Figure S3.**
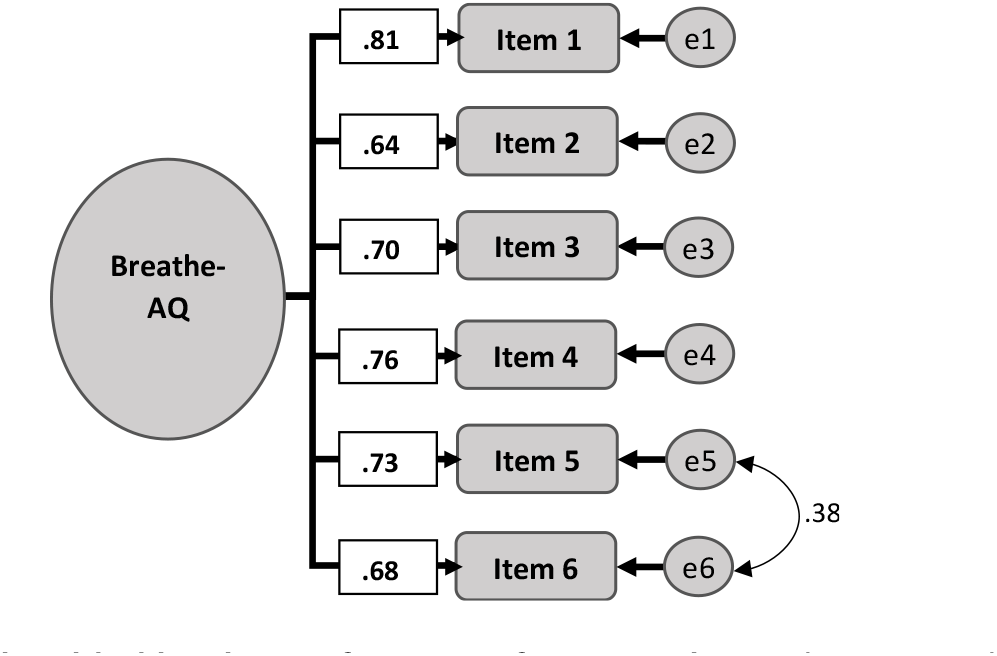
Final overall model yielded by the confirmatory factor analysis. Shown are the standardized item-factor loadings. Abbreviated item numbers refer to the 6 selected items of the Breathing Vigilance Questionnaire (Breathe-VQ). Also shown are the covariance between the residual error terms (‘e’) of items 5 and 6.

**Table S3.**
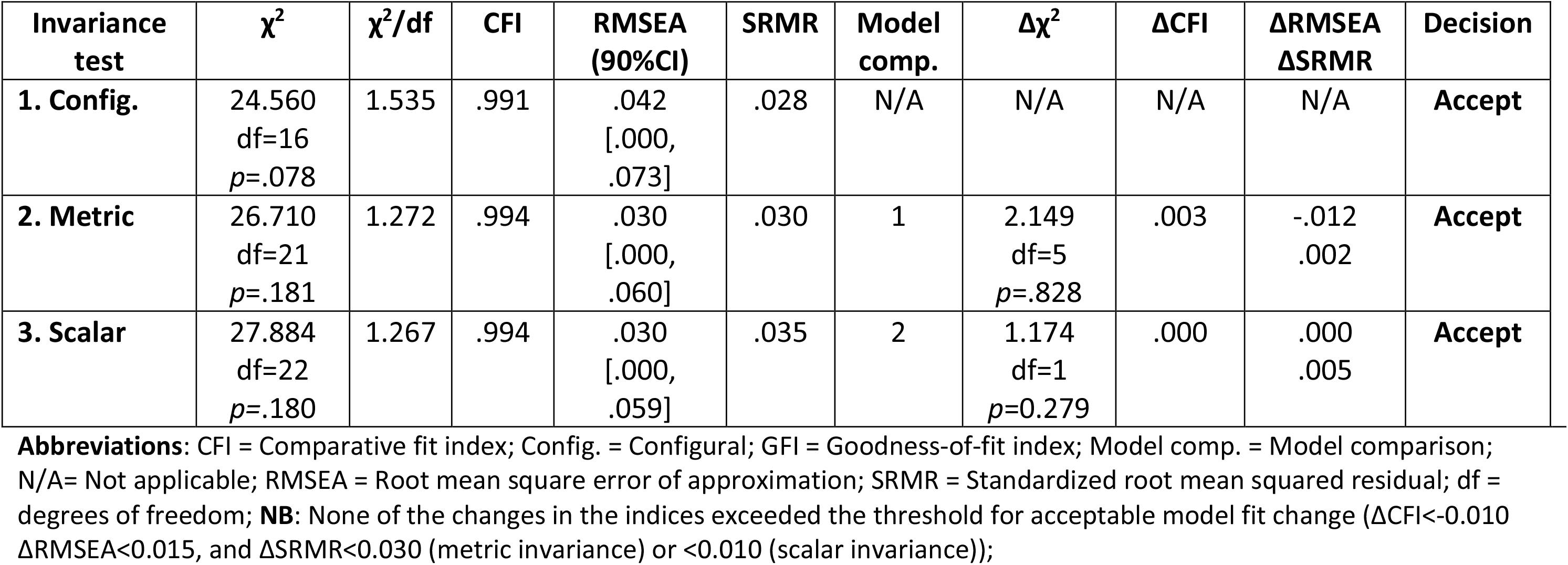
Results of measurement invariance testing.

## Supplemental Material 4. Results of the linear regression analysis

Table S4 presents the results regarding the linear association between breathing vigilance scores (Breathe-VQ) and Nijmegen Questionnaire scores, within a subgroup of people at risk of having DB (N=71). Note that, while 76 participants fell in the ‘high risk of DB’ category, 5 of these could not be included as they had missing items for either the Nijmegen, STAI, or Breathe-VQ questionnaires (and hence scores could not be calculated for these measures).

**Table S4.** Results of regression model.

| **MODEL 1** |  |  |  |  |  |
| --- | --- | --- | --- | --- | --- |
| Dependent variable: **Nijmegen Questionnaire scores (severity of symptoms associated with DB)** |  |  |  |  |  |
|  | *B (SE)* | [95% CI] | *p* | *R*<sup>2</sup> | *R*<sup>2</sup><br>change |
| **Step 1** |  |  |  | .139<br>(*p*=.040) |  |
| Constant | 21.598 (6.678) | [8.265, 34.931] | .002 |  |  |
| Trait Anxiety (STAI) | .206 (.072) | [.062, .350] | .006 |  |  |
| Age (in years) | -.032 (.105) | [-.241, .178] | .763 |  |  |
| Gender | .458 (1.481) | [-2.500, 3.416] | .758 |  |  |
| Depression Diagnosis | -1.013 (1.507) | [-4.021, 1.995] | .504 |  |  |
| **Step 2** |  |  |  | .239<br>(*p*=.003) | .100<br>(.005) |
| Constant | 14.531 (6.773) | [1.005, 28.057] | .036 |  |  |
| Trait Anxiety (STAI) | .203 (.068) | [.066, .339] | .004 |  |  |
| Age (in years) | .018 (.101) | [-.184, .219] | .861 |  |  |
| Gender | .512 (1.403) | [-2.291, 3.315] | .717 |  |  |
| Depression Diagnosis | -1.812 (1.453) | [-4.715, 1.090] | .217 |  |  |
| Breathing Vigilance (Breathe-VQ) | .385 (.132) | [.122, .648] | .005 |  |  |

**Abbreviations:** CI = confidence interval; SE = standard error; STAI = State-Trait Anxiety Inventory;

## Footnotes

1 We excluded people with (ii) or (iii) because we were primarily interested in primary dysfunctional breathing for this initial validation study.

## References

1. Bott J, Blumenthal S, Buxton M, et al. Guidelines for the physiotherapy management of the adult, medical, spontaneously breathing patient. Thorax 2009; 64: 1–52. http://dx.doi.org/10.1136/thx.2008.110726

2. Morgan MDL. Dysfunctional breathing in asthma: is it common, identifiable and correctable? Thorax 2002; 57(Suppl II): ii31–5.

3. Thomas M, McKinley RK, Foy C, et al. Breathing retraining for dysfunctional breathing in asthma: a randomised controlled trial. Thorax 2003; 58: 110–15. http://dx.doi.org/10.1136/thorax.58.2.110

4. Courtney R, Greenwood KM, Cohen M. Relationships between measures of dysfunctional breathing in a population with concerns about their breathing. J Bodywork Mov Ther 2011; 15: 24–34. https://doi.org/10.1016/j.jbmt.2010.06.004

5. Hormbrey J, Jacobi MS, Patil CP, et al. CO2 response and pattern of breathing in patients with symptomatic hyperventilation compared with asthmatic and normal subjects. Eur Respir J 1988; 1: 846–52.

6. Boulding R, Stacey R, Niven R, Fowler SJ. Dysfunctional breathing: a review of the literature and proposal for classification. Eur Resp Rev 2016; 25: 287–94. https://doi.org/10.1183/16000617.0088-2015

7. Hagman C, Janson C, Emtner M. A comparison between patients with dysfunctional breathing and patients with asthma. Clin Resp J 2008; 2: 86–91. https://doi.org/10.1111/j.1752-699X.2007.00036.x

8. Law N, Ruane LE, Low K, et al. Dysfunctional breathing is more frequent in chronic obstructive pulmonary disease than in asthma and in health. Resp Physiol Neurobiol 2018; 247: 20–3. https://doi.org/10.1016/j.resp.2017.08.011

9. Frésard I, Genecand L, Altarelli M, et al. Dysfunctional breathing diagnosed by cardiopulmonary exercise testing in ‘long COVID’ patients with persistent dyspnoea. BMJ Open Resp Res. 2022; 9(1): e001126. https://doi.org/10.1136/bmjresp-2021-001126

10. Thomas M, McKinley RK, Freeman E, et al. The prevalence of dysfunctional breathing in adults in the community with and without asthma. Prim Care Respir J 2005; 14: 78–82. https://doi.org/10.1016/j.pcrj.2004.10.007

11. Ok JM, Park YB, Park YJ. Association of dysfunctional breathing with health-related quality of life: A cross-sectional study in a young population. PLoS One 2018; 13: e0205634. https://doi.org/10.1371/journal.pone.0205634

12. Jones M, Harvey A, Marston L, O’Connell NE. Breathing exercises for dysfunctional breathing/hyperventilation syndrome in adults. Cochrane Database Syst Rev 2013; 5: CD009041. https://doi.org/10.1002/14651858.CD009041.pub2

13. Hagman C, Janson C, Emtner M. Breathing retraining-a five-year follow-up of patients with dysfunctional breathing. Respir Med 2011; 105: 1153–9. https://doi.org/10.1016/j.rmed.2011.03.006

14. Vidotto L, Bigliassi M, Jones M, et al. Stop thinking! I can’t! Do attentional mechanisms underlie primary dysfunctional breathing? Front Physiol 2018; 9 :782. https://doi.org/10.3389/fphys.2018.00782

15. Han JN, Zhu YJ, Li SW, et al. Medically unexplained dyspnea: psychophysiological characteristics and role of breathing therapy. Chin Med J 2004; 117: 6–13. https://doi.org/10.3760/cma.j.issn.0366-6999.2004.01.103

16. Liu S, Ye M, Pao GM, et al. Divergent brainstem opioidergic pathways that coordinate breathing with pain and emotions. Neuron 2022; 110: 857–73. https://doi.org/10.1016/j.neuron.2021.11.029

17. Denton E, Bondarenko J, Tay T, et al. Factors associated with dysfunctional breathing in patients with difficult to treat asthma. J Allergy Clin Immunol Pract 2019; 7: 1471–6. https://doi.org/10.1016/j.jaip.2018.11.037

18. Nardi AE, Freire RC, Zin WA. Panic disorder and control of breathing. Resp Physiol Neurobiol 2009; 167: 133–43. https://doi.org/10.1016/j.resp.2008.07.011

19. Price CJ, Hooven C. Interoceptive awareness skills for emotion regulation: Theory and approach of mindful awareness in body-oriented therapy (MABT). Front Psychol 2018 28; 9: 798. https://doi.org/10.3389/fpsyg.2018.00798

20. Bonaz B, Lane RD, Oshinsky ML, et al. Diseases, disorders, and comorbidities of interoception. Trends Neurosci 2021; 44: 39–51.

21. Schmidt NB, Lerew DR, Trakowski JH. Body vigilance in panic disorder: evaluating attention to bodily perturbations. J Consul Clin Psychol 1997; 65: 214–220. https://psycnet.apa.org/doi/10.1037/0022-006X.65.2.214

22. Harrison OK, Köchli L, Marino S, et al. Interoception of breathing and its relationship with anxiety. Neuron 2021;109: 4080–93. https://doi.org/10.1016/j.neuron.2021.09.045

23. McCracken LM. “Attention” to pain in persons with chronic pain: a behavioral approach. Behav Ther 1997; 28: 271–84. https://doi.org/10.1016/S0005-7894(97)80047-0

24. Kimble M, Boxwala M, Bean W, et al. The impact of hypervigilance: evidence for a forward feedback loop. J Anx Disord 2014; 28: 241–5. https://doi.org/10.1016/j.janxdis.2013.12.006

25. Popkirov S, Staab JP, Stone J. Persistent postural-perceptual dizziness (PPPD): a common, characteristic and treatable cause of chronic dizziness. Practical Neurol 2018; 18: 5–13. http://dx.doi.org/10.1136/practneurol-2017-001809

26. Ellmers TJ, Kal EC, Young WR. Consciously processing balance leads to distorted perceptions of instability in older adults. J Neurol 2021; 268: 1374–84. https://doi.org/10.1007/s00415-020-10288-6

27. De Peuter S, Janssens T, Van Diest I, et al. Dyspnea-related anxiety: the Dutch version of the Breathlessness Beliefs Questionnaire. Chron Resp Dis 2011; 8: 11–9. https://doi.org/10.1177%2F1479972310383592

28. Banzett RB, O’Donnell CR, Guilfoyle TE, et al. Multidimensional Dyspnea Profile: an instrument for clinical and laboratory research. Eur Resp J 2015; 45: 1681–91. http://dx.doi.org/10.1183/09031936.00030115

29. Yorke J, Moosavi SH, Shuldham C, Jones PW. Quantification of dyspnoea using descriptors: development and initial testing of the Dyspnoea-12. Thorax 2010; 65: 21–6. http://dx.doi.org/10.1136/thx.2009.118521

30. Mehling WE, Acree M, Stewart A, et al. The multidimensional assessment of interoceptive awareness, version 2 (MAIA-2). PloS One 2018; 13: e0208034. https://doi.org/10.1371/journal.pone.0208034

31. Hollins M, Harper D, Gallagher S, et al. Perceived intensity and unpleasantness of cutaneous and auditory stimuli: an evaluation of the generalized hypervigilance hypothesis. Pain 2009; 141: 215–21. https://doi.org/10.1016/j.pain.2008.10.003

32. Field, A., Discovering statistics using IBM SPSS statistics. 5th Edn. Sage Publications Ltd, London, 2018.

33. Heathcote LC, Simons LE. Stuck on pain? Assessing children’s vigilance and awareness of pain sensations. Eur J Pain 2020; 24: 1339–47. https://doi.org/10.1002/ejp.1581

34. Van Dixhoorn J, Duivenvoorden HJ. Efficacy of Nijmegen Questionnaire in recognition of the hyperventilation syndrome. J Psychosom Res 1985; 29: 199–206. https://doi.org/10.1016/0022-3999(85)90042-X

35. Spielberger CD, Gorsuch RL, Lushene R, et al. Manual for the State-Trait Anxiety Inventory. Palo Alto, CA: Consulting Psychologists Press, 1983.

36. Masters RSW, Eves FF, Maxwell J. Development of a movement specific Reinvestment Scale. Proceedings of the ISSP 11th World Congress of Sport Psychology, Sydney, Australia, 2005.

37. Portney LG, Watkins MP. Foundations of clinical research: applications to practice. 3rd Edn. Upper Saddle River, NJ, Pearson/Prentice Hall, 2009.

38. Stevens JP. Applied multivariate statistics for the social sciences. 4th Edn. Hillsdale, New York: Erlbaum, 2002.

39. West SG, Finch JF, Curran PJ. Structural equation models with non-normal variables. Problems and remedies. In: Hoyle RH, ed. Structural equation modeling: concepts, issues and applications. Newbury Park, CA: Sage; 1995; pp. 56–75.

40. Hu L, Bentler PM. Cut-off criteria for fit indexes in covariance structure analysis: conventional criteria versus new alternatives. Struct Equ Modeling 1999; 6: 1–55. https://doi.org/10.1080/10705519909540118

41. Medsker GJ, Williams LJ, Holahan PJ. A review of current practices for evaluating causal models in organizational behavior and human resources management research. J Manage 1994; 20: 439–64. https://doi.org/10.1177%2F014920639402000207

42. Browne MW, Cudeck R. Alternative ways of assessing model fit. Sociol Method Res 1992; 21: 230–58. https://doi.org/10.1177%2F0049124192021002005

43. Chen FF. Sensitivity of goodness of fit indexes to lack of measurement invariance. Struct Equ Modeling 2007; 14: 464–504. https://doi.org/10.1080/10705510701301834

44. Weir JP. Quantifying test-retest reliability using the intraclass correlation coefficient and the SEM. J Strength Cond Res 2005; 19: 231–40. https://doi.org/10.1519/15184.1

45. De Vet HCW, Terwee CB, Knol DL, et al. When to use agreement versus reliability measures. J Clin Epidemiol 2006; 59: 1033–9. https://doi.org/10.1016/j.jclinepi.2005.10.015

46. Mokkink LB, Terwee CB, Knol DL, et al. The COSMIN checklist for evaluating the methodological quality of studies on measurement properties: a clarification of its content. BMC Med Res Methodol 2010; 10: 22. https://doi.org/10.1186/1471-2288-10-2

47. Streiner DL, Norman GR. Health measurement scales. A practical guide to their development and use. 4th Ddn. New York: Oxford University Press, 2008.

48. Ellmers TJ, Wilson MR, Kal EC, Young WR. Standing up to threats: Translating the two-system model of fear to balance control in older adults. Exp Gerontol 2022; 158: 111647. https://doi.org/10.1016/j.exger.2021.111647

49. Weng HY, Feldman JL, Leggio L, Napadow V, Park J, Price CJ. Interventions and manipulations of interoception. Trends Neurosci 2021; 44: 52–62. https://doi.org/10.1016/j.tins.2020.09.010

50. Lewis A, Cave P, Stern M, et al. Singing for Lung Health—a systematic review of the literature and consensus statement. NPJ Prim Care Respir Med 2016; 26: 16080. https://doi.org/10.1038/npjpcrm.2016.80

51. Todd S, Walsted ES, Grillo L, Livingston R, Menzies-Gow A, Hull JH. Novel assessment tool to detect breathing pattern disorder in patients with refractory asthma. Respirology. 2018; 23: 284–90. https://doi.org/10.1111/resp.13173

